# Subclinical Atherosclerosis Links Premature and Late-Onset Coronary Artery Disease

**DOI:** 10.1101/2025.11.06.25339722

**Authors:** Miao Guo, Mantong Zhao, Yanchen Zhao, Anxin Wang, Xiuhua Guo, Lixin Tao, Jia Liu

## Abstract

**Background:** Despite established relationship between atherosclerosis and coronary artery disease (CAD), evidence on subclinical atherosclerosis and its differential associations with premature coronary artery disease (PCAD) versus late-onset coronary artery disease (LCAD) remains limited.

**Aims:** This study aims to delve deeper into the associations between subclinical atherosclerosis and the incidence risk of both PCAD and LCAD.

**Methods:** Using UK Biobank data, we identified PCAD (male <55/female <65 years; n=7,398) and LCAD (male ≥55/female ≥65 years; n=39,085) cohorts. Conditional inference tree classification optimized carotid intima-media thickness (cIMT) stratification in both cohorts.

**Results:** Conditional inference tree categorized the PCAD cohort into two subgroups: cIMT ≤700μm and cIMT >700μm, with the latter demonstrating a HR of 2.079 for cardiovascular risk. In the LCAD cohort, four cIMT strata were identified: ≤ 620μm, 620–763μm (HR=1.401), 763–1054μm (HR=1.810), and >1054μm (HR=2.850). Multivariable-adjusted Cox models demonstrated significant associations between subclinical atherosclerosis and PCAD (HR=2.079, 95%CI:1.477-2.925) and LCAD (HR=1.776, 95%CI:1.455-2.169), highlighting the prognostic value of cIMT stratification in coronary artery disease risk assessment.

**Conclusions:** A UK Biobank prospective cohort study revealed subclinical atherosclerosis significantly associated with PCAD and LCAD risks. Even within conventional cIMT “safe thresholds”, incremental increases predicted elevated risks, underscoring limitations of current thresholds. Future research need to develop multimodal frameworks integrating dynamic cIMT trajectories to refine risk stratification and early interventions.

## 1. Introduction

Premature coronary artery disease (PCAD), a distinct subset of coronary artery disease (CAD), is characterized by its onset in individuals aged 55 years or younger for males and 65 years or younger for females*[1]*. This type of CAD is marked by significant coronary atherosclerosis, featuring a high burden of non-calcified plaques that dominate the pathology. Additionally, PCAD patients exhibit notable high-risk characteristics such as spotty calcification, positive remodeling, and low attenuation*[2]*, which collectively contribute to a heightened risk of ischemic recurrence and premature mortality*[3]*. Meta-analyses suggest potential differences in predictive validity and interaction patterns of multiple risk factors between PCAD and later-onset coronary artery disease (LCAD)*[4]*.

In clinical practice, the distinction between atherosclerosis and subclinical atherosclerosis primarily hinges on the presence or absence of clinical ischemic symptoms, such as stroke, myocardial infarction, or chronic limb ischemia[5]. The concept of subclinical atherosclerosis stems from the recognition of atherosclerotic lesions in asymptomatic patients, often incidentally detected through non-invasive imaging techniques such as carotid or femoral artery imaging, echocardiography, or coronary computed tomography (CT). As the initial phase of atherosclerosis, subclinical atherosclerosis has not yet resulted in significant stenosis of vital arteries or overt clinical manifestations. Consequently, the presence of subclinical atherosclerosis offers a valuable window of opportunity for the prevention and early intervention of coronary atherosclerotic heart disease. Enhancing early diagnosis and effective treatment of subclinical atherosclerosis is regarded as a critical turning point in halting the progression to CAD, enabling the implementation of early, multimodal, and innovative preventive strategies in primary and secondary prevention for younger patients with CAD.

In 2005, Espeland et al. conducted an in-depth comprehensive analysis of seven clinical trials on statin drugs and, for the first time, established the thickening of carotid intima-media thickness (cIMT) as an important surrogate marker for assessing CVD risk*[6]*. Subsequently, the increase in cIMT has been widely recognized as the most sensitive and reliable biomarker for the early stage of subclinical atherosclerosis, reflecting early abnormalities in arterial wall structure and function[7].

Although previous studies have investigated the association between subclinical atherosclerosis and CAD[8–11], evidence on the independent relationship of subclinical atherosclerosis burden with both PCAD and LCAD risks remains largely absent in the existing literature. Furthermore, current strategies for cardiovascular risk stratification in complex clinical networks remain suboptimal, particularly in addressing the interplay between subclinical atherosclerosis and age-dependent diseases. To address this gap, we first leveraged conditional inference tree (CTree) methodology to refine cIMT stratification, enabling dynamic partitioning of atherosclerosis burden across heterogeneous populations. Subsequently, using UK Biobank data, we conducted multivariable-adjusted Cox proportional hazards regression analyses to evaluate the associations between cIMT strata and incident PCAD and LCAD. Finally, to ensure analytical rigor, we implemented a comprehensive validation framework incorporating multi-angle cIMT measurements.

## 2. Methods

### 2.1. Study population and design

The UK Biobank represents an extensive prospective cohort study, encompassing over half a million participants aged 37 to 73, recruited across England, Scotland, and Wales from 2006 to 2010*[12]*. UK Biobank has amassed a wealth of data, spanning Participant Information, Population Characteristics, Assessment Center records, Biological Samples, Genomics insights, Online Follow-ups, Additional Exposures, and Health-related Outcomes—all meticulously documenting longitudinal health trends.

To investigate the complex relationship between mean cIMT and the incidence of both PCAD and LCAD, we conducted a longitudinal cohort study utilizing follow-up data from the UK Biobank in 2014.

From the initial 502,152 UK Biobank participants, 453,060 with missing cIMT measurements, 94 declining core questionnaire responses, and 2,514 with baseline coronary artery disease were excluded, yielding 46,483 eligible participants for analysis, showed in figure 1. For the PCAD analysis, 7,398 participants were included, defined as those diagnosed with PCAD before age <55 years (male)/<65 years (female) or reaching the end of follow-up without exceeding these age thresholds, whichever occurred first. For the LCAD analysis, 39,085 participants were included, those diagnosed with PCAD before age ≥55 years (male)/ ≥65 years (female) or reaching the end of follow-up without exceeding these age thresholds, whichever occurred first. In the PCAD analysis, 162 incident PCAD cases (2.19%) were recorded over a median follow-up of 8.9 years, with 7,236 participants (97.81%) completing follow-up without developing PCAD. In the LCAD analysis, 1,473 incident LCAD cases (3.77%) were recorded over a median follow-up of 8.9 years, with 37,612 participants (96.23%) completing follow-up without developing LCAD.

**Figure 1.**
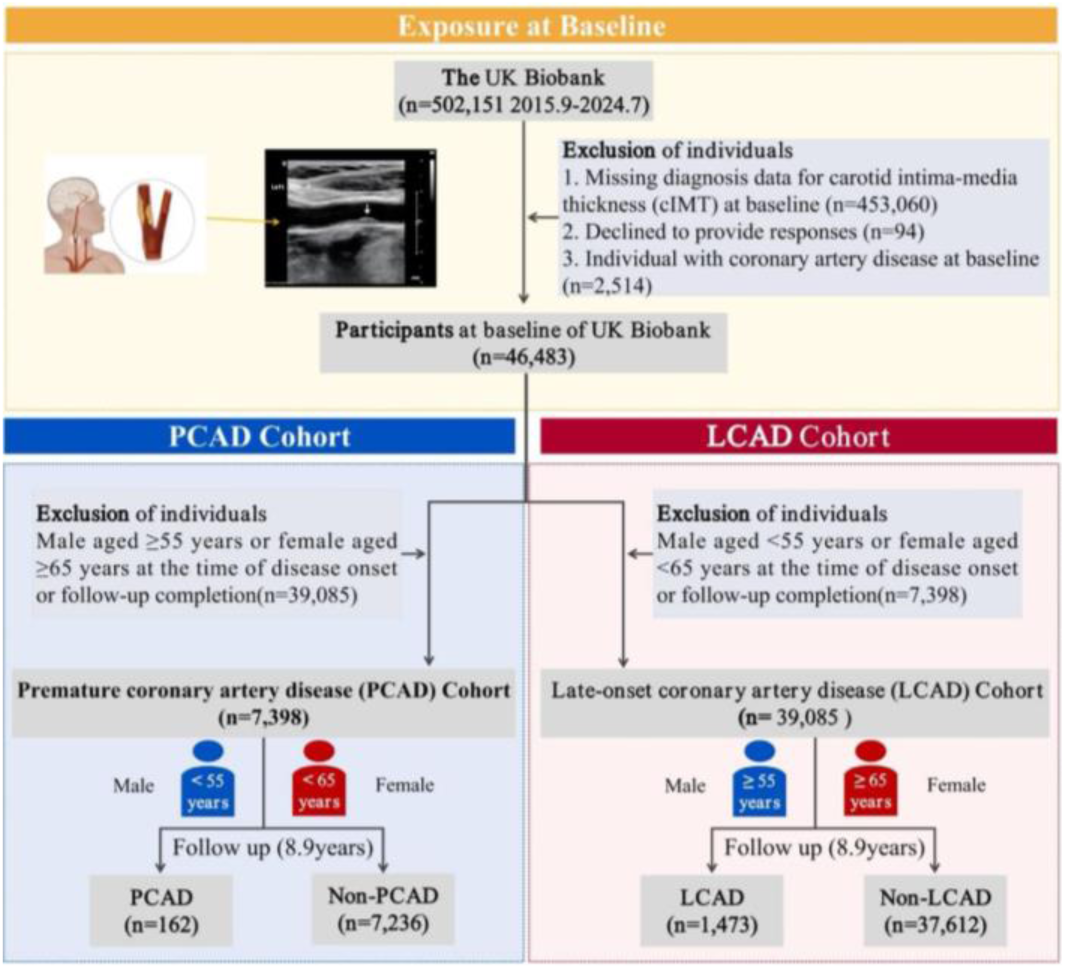
Flowchart of the study population

### 2.2. cIMT Measurements

At the time of the analysis, measurements of mean cIMT were available for 50,026 participants who had completed the third follow-up of the UK Biobank study. These mean cIMT measurements were automatically acquired from 2D carotid artery scans utilizing a cardiac health station supplied by Panasonic Biomedical Sales Europe BV, located in Leicestershire, United Kingdom. A comprehensive total of four mean cIMT measurements were conducted, specifically at angles of 120° and 150° in the right carotid artery, and 210° and 240° in the left carotid artery.*[13]*

Quality control of cIMT measurements was performed by the UK Biobank, validated both internally and externally using a set of predefined criteria[14]. CIMT measurements that did not pass the quality control, either with cIMT values of zero or as indicated by the corresponding quality control flags (field IDs 22682, 22683, 22684, 22685) were excluded from the analyses*[15]*. Measurements collected at the first (imaging visit) were considered for the current analysis. The mean cIMT at 120 degrees was taken as the final cIMT measurement.

According to the Mannheim consensus, plaque was defined as any focal thickening or protruding into the lumen with cIMT > 1.5mm thickness, and anterior plaque was defined as any focal thickening with 1.0mm <cIMT≤1.5mm*[16]*. Therefore, due to the small proportion of people with cIMT greater than 1.0mm, in order to optimize the effectiveness of risk stratification, we adopted a dichotomized classification of mean cIMT: <1.00mm as normal (45029/46483) and ≥1.0mm (1454/46483) as subclinical atherosclerosis.

### 2.3. Ascertainment of outcomes (2015.9-2024.8)

The diagnosis of CAD in the UK Biobank’s “Health-related Outcomes - First Incidence - Circulatory System Disorders” category was established based on self-reported information. Participants were instructed to disclose all physician-diagnosed health conditions during their recruitment visit to the assessment clinic. Subsequently, their responses were corroborated by nurses through an oral interview. Specifically, the following three conditions were defined as CAD outcomes in accordance with the International Statistical Classification of Diseases and Related Health Problems, angina pectoris (ICD-10 code: I20), acute myocardial infarction (ICD-10 code: I21), and chronic ischemic heart disease (ICD-10 code: I25).

### 2.4. Covariates

Age and sex were obtained by UK Biobank staff prior to an assessment visit at the local NHS primary care trust registry. Systolic blood pressures (SBP) and diastolic blood pressures (DBP), as well as body mass index (BMI), were assessed by UK Biobank staff at the assessment center. Among automatic and manual measurements, automatic measurement was preferred for blood pressure assessment. BMI was calculated by dividing weight by the square of height. The UK Biobank utilized the Tanita BC-418 AM body composition analyzer to measure weight and the Seca 202 height gauge to measure height*[15]*. Participants were instructed by UK Biobank staff to remove heavy clothing and shoes and to stand on the foot pads of the body composition analyzer. Smoking status, alcohol drinker status, diabetes, educational qualifications, duration of activity, average total household income before tax, father’s and mother’s family history of heart disease were collected via a touch-screen questionnaire during the assessment visit.

In the current study, participants were classified into three categories: “never smokers/drinkers,” “former smokers/drinkers,” and “current smokers/drinkers,” based on their responses to touchscreen questionnaires regarding their smoking and alcohol consumption status. Furthermore, participants were dichotomized into those with higher than a bachelor’s degree and those with lower than a bachelor’s degree, according to their responses to the touchscreen questionnaire on educational attainment. Average total household income before tax was categorized into the following brackets: “Less than £18,000,” “£18,000 to £30,999,” “£31,000 to £51,999,” “£52,000 to £100,000,” and “Greater than £100,000.” Other categorical variables were classified in accordance with the criteria established by the UK Biobank.

Using WHO BMI classification criteria, obesity was defined as BMI >30 kg/m², with remaining participants categorized as normal*[17]*. Hypertension was operationally defined as SBP ≥140 mmHg and/or DBP ≥90 mmHg, consistent with ESC hypertension diagnostic guidelines*[18]*.

### 2.5. Statistical analysis

This study utilized data from the UK Biobank cohort. Follow-up time for each participant was calculated from September 1, 2015, to July 31, 2024, defined as the period from the baseline survey date to the date of CAD diagnosis or the end of follow-up, whichever occurred first. Incidence rates of PCAD and LCAD were calculated based on the proportion of incident cases within the cohort during the follow-up period. To examine the relationship between cIMT and PCAD/LCAD, statistical analyses were performed using R.

All prospective analyses were adjusted for sex, age, BMI, alcohol drinker status, smoking status, diabetes diagnosed by doctor, SBP, DBP, total vigorous physical activity time, average household income after tax, education level and family history of heart disease, and were stratified by cIMT. Missing covariate data were imputed using the Multiple Imputation by “MICE” package*[19]*.

We employed Conditional inference tree (CTree) to precisely determine the optimal partitioning scheme for cIMT*[20]*. In this process, the continuous variable cIMT can be repeatedly split at different nodes of the CTree. Each splitting step follows formal statistical inference, quantifying covariate-outcome associations via regression models. A split node is selected only when the global null hypothesis, which assumes no covariate has a univariate association with the outcome, is rejected. The covariate with the strongest association is chosen for splitting. CTree controls tree size using a single type I error rate parameter, α, eliminating pruning needs. The α value is adjustable independently of outcome type, such as continuous, binary, or event time. CTree’s complexity parameter is independent of splitting criteria, ensuring consistent tree-building. Through multiplicity-adjusted inference, CTree provides statistical guarantees and valid p-values at each step, ensuring model robustness*[21]*. Through this recursive process, we sequentially determined the numerical range for each interval and calculated the number of individuals with and without the outcome, as well as the percentage of individuals with the outcome in each group. For the LCAD dataset, cIMT was categorized using the CTree method. However, due to data limitations, quartile-based classification was applied to the PCAD dataset.

To investigate the relationship between varying degrees of cIMT and PCAD/LCAD events, cIMT was treated as a binary variable, a CTree-derived categorical variable, or one standard deviation of cIMT. We analyzed the association of each classification with the incidence of coronary artery disease (including angina, acute myocardial infarction, and chronic ischemic heart disease) during the study period. Additionally, based on Cox regression analysis results, we constructed Kaplan-Meier survival curves, restricted cubic spline (RCS) curves, and forest plots to visually describe these associations.

### 2.6. Sensitivity analysis

To reverify the viability of cohort findings, sensitivity analyses were conducted focusing on cIMT measurement angles: cIMT data from four angles (right 120°, 150°; left 210°, 240°) were acquired via standardized ultrasound protocols. The 120° measurement was selected as the primary analytical variable. Participants were stratified into PCAD (cIMT≥1.0mm) and normal groups (cIMT<1.0mm)*[16]*. CTree algorithm dynamically categorized cIMT values across angles, with multivariable Cox proportional hazards models assessing associations with PCAD.

We conducted subgroup analyses to evaluate the relationship between cIMT and PCAD and LCAD, stratified by sex (female, male), family history of heart disease (yes,no), obesity (yes, no), hypertension (yes, no), diabetes (yes, no), smoking status (never smoked, former smoker, current smoker), education level (university degree or above, below university degree), and household income (less than £18,000, £18,000 to £30,999, £31,000 to £51,999, £52,000 to £100,000, greater than £100,000).

## 3. Results

### 3.1. Association between cIMT and PCAD

This study analyzed 7,398 adults in the PCAD cohort and 39,085 adults in the LCAD cohort. Among these, 162 PCAD cases were identified in the premature cohort, and 1,473 LCAD cases were identified in the late-onset cohort. Participants were grouped based on CAD incidence at the end of follow-up. Other variables, including sex, age, BMI, alcohol drinker status, smoking status, diabetes diagnosed by doctor, SBP, DBP, total vigorous physical activity time, average household income after tax, education level and family history of heart disease showed statistically significant differences (P<0.05), as detailed in Table 1. Compared to non-PCAD individuals, those with PCAD exhibited higher cIMT and prevalence of diabetes, obesity, hypertension and elevated education levels, LCAD cases showed the same as PCAD.

Compared to non-PCAD individuals, PCAD cases demonstrated significantly higher cIMT, prevalence rates of diabetes, obesity, hypertension, and elevated educational attainment. LCAD cases exhibited parallel patterns to PCAD (all P<0.01), as detailed in Supplementary Table 1.

Subclinical atherosclerosis, a critical pathological substrate underlying the global burden of CAD22, requires further elucidation regarding its association with PCAD. Our analysis revealed that compared to the lowest group (0-700μm), the unadjusted HR for the elevated cIMT group (700-1496μm) was 2.570 (95%CI: 1.841-3.586, P<0.001). After multivariable adjustment (sex, age, BMI, alcohol intake, smoking, diabetes, blood pressure, physical activity, socioeconomic status, and family medical history), the association remained significant (HR=2.079, 95% CI: 1.477-2.925, P<0.001). RCS analysis demonstrated a notable linear relationship between cIMT and PCAD risk (nonlinearity P=0.431; overall P=0.001), as illustrated in Figure 2.

**Figure 2.**
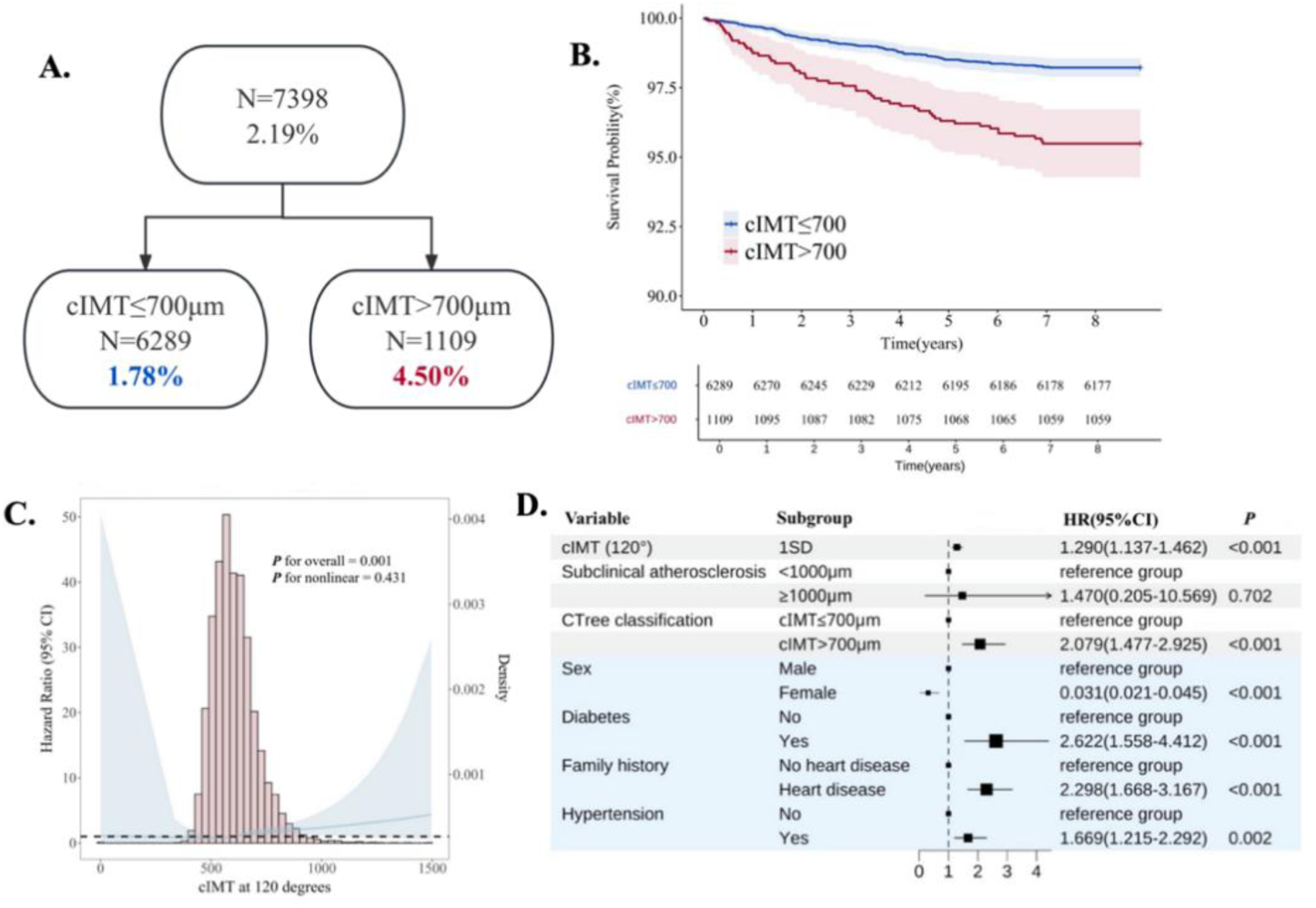
Relationship between subclinical atherosclerosis and premature coronary artery disease. (A) Conditional inference tree algorithm for stratifying participants into four distinct risk groups based on carotid intima-media thickness thresholds. (B, C, D) Assessment of relative risk for all-cause mortality in four cIMT groups. (B) Kaplan-Meier survival curves showing time-to-late-onset coronary artery disease analysis across cIMT groups over follow-up: Cox proportional hazards model (adjusted for sex, age, obesity, alcohol consumption, smoking, diabetes, hypertension, physical activity, household income, education, and family history of heart disease). (C) Hazard ratios (solid line; same multivariable-adjusted Cox model as B) with 95% CIs (blue shading) show premature coronary artery disease risk by cIMT levels, modeled via restricted cubic splines. Right y-axis density plots reflect baseline cIMT distribution (standardized, positively skewed) (D) Forest plot with hazard ratios for coronary events across cIMT-based risk strata in the premature coronary artery disease cohort.

### 3.2. Association between cIMT and LCAD

To assess the association between cIMT and LCAD among participants, we conducted a time-to-event analysis focusing onLCAD (Figure 3). Initially, we applied the CTree classification to stratify study participants into four groups based on their cumulative all-cause mortality rates at the end of follow-up. The group with cIMT ranging from 0 to 620 μm exhibited the lowest LCAD incidence (2.61%, 336/12,878 cases). The LCAD incidence increased in the 620 to 763 μm group (3.62%, 551/15,227 cases) and the 763 to 1054 μm group (5.05%, 511/10,110 cases). The highest LCAD incidence was observed in the 1054 to 1971μm group (8.62%, 75/870 cases). The log-rank test yielded similar results, indicating a continuous increase in the incidence of LCAD with increasing cIMT.

**Figure 3.**
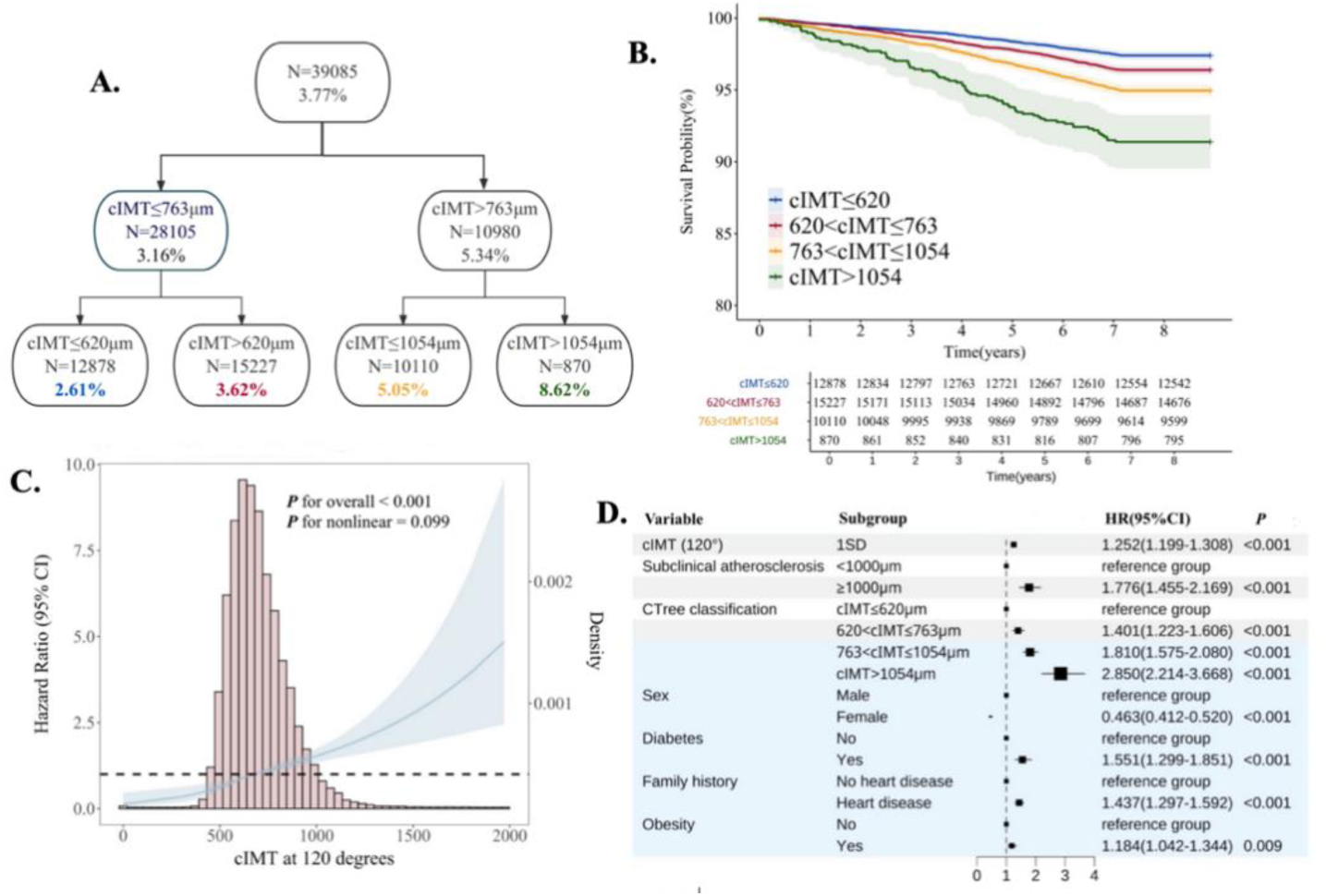
Relationship between subclinical atherosclerosis and late-onset coronary artery disease. (A) Conditional inference tree algorithm for stratifying participants into four distinct risk groups based on carotid intima-media thickness thresholds. (B,C,D) Assessment of relative risk for all-cause mortality in four cIMT groups. (B) Kaplan-Meier survival curves showing time-to-late-onset coronary artery disease analysis across cIMT groups over follow-up: Cox proportional hazards model (adjusted for sex, age, obesity, alcohol consumption, smoking, diabetes, hypertension, physical activity, household income, education, and family history of heart disease). (C) Hazard ratios (solid line; same multivariable-adjusted Cox model as B) with 95% CIs (blue shading) show late-onset coronary artery disease risk by cIMT levels, modeled via restricted cubic splines. Right y-axis density plots reflect baseline cIMT distribution (standardized, positively skewed) (D) Forest plot with hazard ratios for coronary events across cIMT-based risk strata in the late-onset coronary artery disease cohort.

Without any adjustment for confounding factors, the risk of LCAD increased by 39.4%, 96.2%, and 242.1% in the 620-763μm, 763-1054μm, and 1054-1971μm groups, respectively, compared to the 0-620 μm group based on cIMT. After adjusting for potential confounding factors such as age, sex, obesity, alcohol drinker status, smoking status, diabetes diagnosed by doctor, hypertension, total vigorous physical activity time, average household income after tax and education level and family history of heart disease using Cox proportional hazards model for time-to-event analysis, the risks of LCAD increased by 40.1%, 81.0%, and 185.0% in the 620-763μm, 763-1054μm, and 1054-1971μm groups, respectively, compared to the 0-620μm group. Additionally, in the RCS analysis with the mean cIMT index as a constant variable, a significant non-linear relationship was observed between this index and LCAD. Furthermore, the model indicated linear relationship between cIMT and LCAD (P for overall<0.001, P for nonlinear=0.099).

### 3.3. Sensitivity analysis

Figure 4A presents the sensitivity analyses evaluating cIMT measurements at various angles, consistently demonstrating significant associations with the risk of PCAD. HRs remained statistically significant for all angles and were highly concordant with the primary analyses, confirming the robustness of our findings to angle selection. Subclinical atherosclerosis at 210° and 240° in the left carotid artery exhibited a stronger association with PCAD risk compared with 120° and 150° in the right carotid artery. This observation suggests that higher cIMT measurement angles may be associated with an increased risk of PCAD. Moreover, it highlights a potential heterogeneity in the impact of carotid intima-media thickening on early-onset coronary artery disease between the left and right carotid arteries, as presented in Supplementary Figure 1, 2, and 3. In contrast, while Figure 4B demonstrates consistent associations between cIMT measurements and LCAD risk across all angles, no significant positive relationship with increasing angles was observed, as presented in Supplementary Figure 4, 5, and 6, highlighting distinct pathological trajectories between early- and late-onset disease phenotypes.

**Figure 4.**
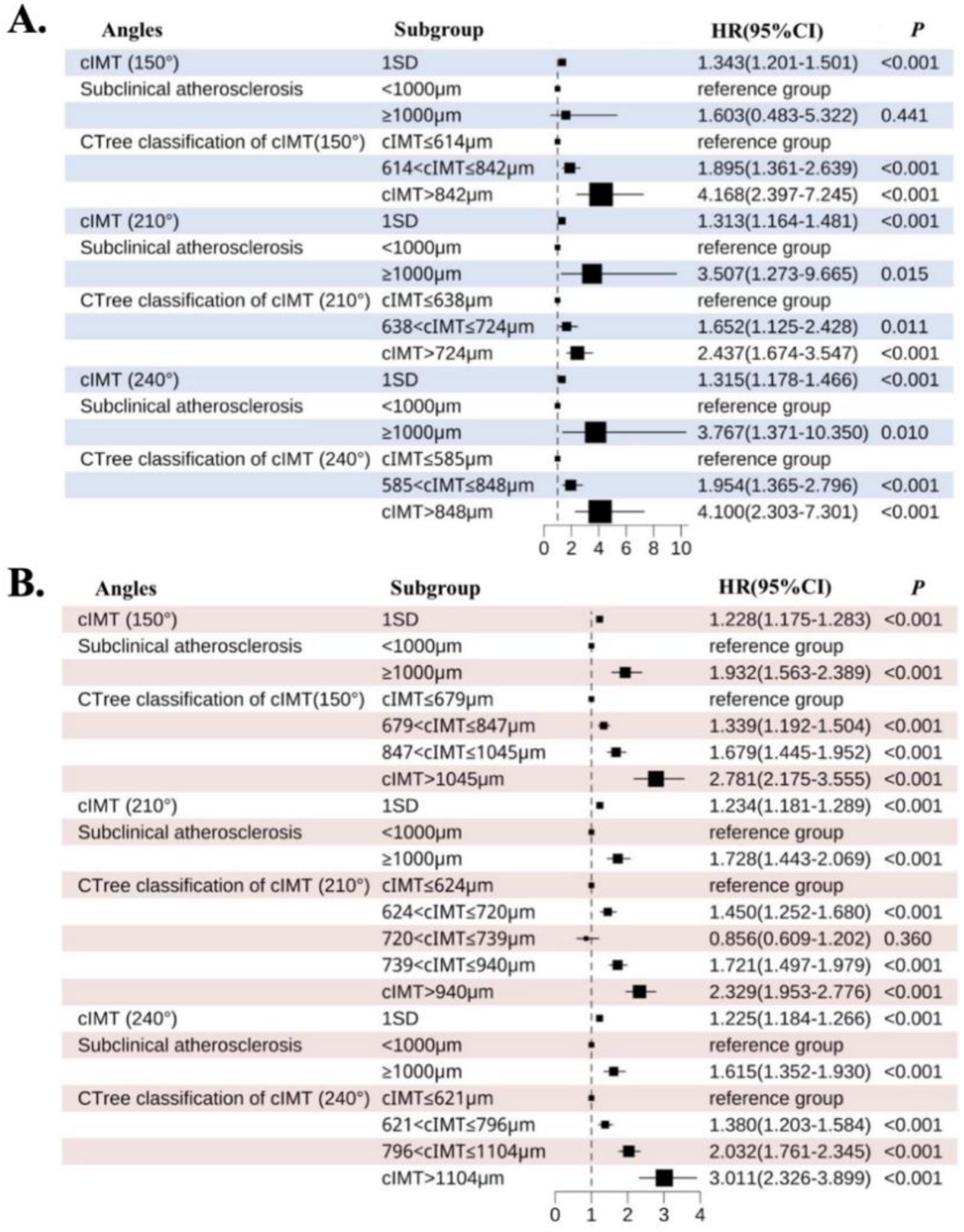
Analysis of cIMT measurement angles: Cox proportional hazards model (adjusted for sex, age, obesity, alcohol consumption, smoking, diabetes, hypertension, physical activity, household income, education, and family history of heart disease). (A) Forest plot with hazard ratios for coronary events across different angles of cIMT-based risk strata in the premature coronary artery disease cohort. (B) Forest plot with hazard ratios for coronary events across different angles of cIMT-based risk strata in the late-onset coronary artery disease cohort.

The results of the subgroup analyses, as presented in Supplementary Table 3 and 4, demonstrate a consistent association between cIMT and both PCAD and LCAD among individuals with obesity, hypertension, and diabetes mellitus. Furthermore, no significant interactions were observed between cIMT and the stratifying variables.

## 4. Discussion

There has been growing academic interest in elucidating the heterogeneity of risk factors between PCAD (men <55 years, women <65 years) and LCAD (men ≥55 years, women ≥65 years). Although CAD predominantly affects older populations, the disease burden and unique pathophysiological characteristics in younger patients warrant attention.

This study leveraged the prospective UK Biobank cohort (median follow-up 8.9 years) to systematically investigate the association patterns between subclinical atherosclerosis and CAD across distinct age-at-onset windows (premature: men <55 years, women <65 years; late-onset: men ≥55 years, women ≥65 years). After multivariable adjustment for conventional cardiovascular risk factors, cIMT stratification exhibited a linear positive association with both PCAD and LCAD risks. The highest cIMT-CTree group demonstrated 2.08-fold elevated risk versus the reference, indicating that subclinical atherosclerotic burden amplifies PCAD risk through accelerated vascular remodeling and endothelial dysfunction. These findings necessitate dynamic cIMT surveillance frameworks to overcome limitations of static threshold-based risk models.

### 4.1. Comparison with previous studies

Multiple large-scale cohort studies have suggested a potential association between an increase in cIMT due to subclinical atherosclerosis and the risk of CAD[10,16,23]. A cIMT >1.0 mm is consistently identified as a high-risk threshold across all ages[24]. Its quantifiability and reproducibility enhance its utility in assessing arterial wall thickening and plaque formation, with strong predictive value in longitudinal analyses*[25]*. The US ARIC study demonstrated that incorporating cIMT and plaques improved CAD risk prediction beyond traditional factors (ΔAUC=0.013; NRI=9.9%)*[26]*, highlighting the potential clinical value of cIMT in optimizing existing risk assessment models. Nair et al. reported an 11% CAD risk increase per 0.1 mm rise in cIMT*[27]*. Another ARIC analysis revealed a two-fold CAD risk in individuals with cIMT >1 mm, with the highest quartile showing a 36% increased risk per 0.19 mm increment*[28]*. The Rotterdam Study confirmed its independent association with CAD events in those ≥55 years*[29]*. Despite these findings, the role of subclinical atherosclerosis in PCAD remains poorly defined. Using UK Biobank data, we identified a linear relationship between cIMT and both PCAD and LCAD, with stronger effects on LCAD. Notably, risk increases were evident even below the 1.0 mm threshold, suggesting a need to re-evaluate current cIMT cutoffs and further explore mechanistic pathways, especially in younger populations.

By phenotypically stratifying CAD into PCAD and LCAD subtypes, this study demonstrated independent associations between cIMT gradients and both entities. RCS analysis revealed linear dose-response relationships, systematically establishing subclinical atherosclerosis as a pathogenic driver of PCAD. Crucially, the 10-year earlier median CAD onset in South Asians (53 years) versus Europeans (63 years)[30–32]. utilizes this population as a unique natural model for delineating atherosclerosis spatiotemporal heterogeneity. Beyond cIMT’s independent effect, multivariable Cox models identified traditional risk factors such as hypertension, diabetes[33–38], and family history of heart disease[39] were significantly linked to PCAD*[40]*. These associations, expressed through distinct analytical frameworks, align well with previous studies.

Furthermore, the Rotterdam Study established cIMT as an independent predictor of cardiovascular events in individuals aged ≥55 years*[29]*. By defining LCAD through internationally standardized sex-specific age thresholds (male ≥55/female ≥65 years), this study provides the cross-ethnic validation of this association, demonstrating the pan-population robustness of cIMT-cardiovascular risk relationships. By optimizing risk stratification through CTree classification, the analytical precision, model robustness, and clinical applicability of our findings were significantly enhanced. The results revealed a positive correlation between increasing cIMT and LCAD risk. Multivariable Cox proportional hazards analyses demonstrated persistent associations, with the 620-763μm, 763-1054μm, and 1054-1971μm cIMT groups exhibiting 40.1%, 81.0%, and 185.0% elevated risks of LCAD, respectively, compared to the reference group (0–620μm). These findings are highly consistent with conclusions from the Rotterdam cohort. Independent of cIMT effects, multivariable Cox proportional hazards models revealed significant associations between LCAD risk and obesity, diabetes and family history of heart disease which was consistent with previous studies*[41]*.

### 4.2. Advantages of this research

This study provides a novel public health perspective on atherosclerotic health disparities. Systematic analysis of a large-scale prospective cohort revealed cIMT, an imaging biomarker of subclinical atherosclerosis, to exhibit independent associations with both PCAD and LCAD. To our knowledge, this is the first large-scale cohort study to systematically investigate the association between subclinical atherosclerosis, assessed by cIMT, and both PCAD and LCAD across distinct clinical onset stages. Previous research has predominantly focused on CAD as a whole, whereas our study provides novel insights into the differential impact of cIMT across early and late disease presentations, offering a more nuanced understanding of atherosclerosis progression in relation to CAD risk. Furthermore, employing the CTree methodology to stratify risk in individuals with cIMT below conventional thresholds, we identified residual CAD risk even within the “safe” cIMT range. The CTree approach demonstrated strengths in dynamic classification, capturing nonlinear relationships, and identifying high-risk subgroups. Finally, by employing multi-angle carotid assessments (yielding consistent findings across four anatomical planes) alongside subgroup analyses to affirm both statistical robustness and external applicability, we have substantiated the strong linkage between subclinical atherosclerosis and PCAD. The correlation for subclinical atherosclerosis at 210° and 240° in the left carotid artery was considerably higher than that at 120° and 150° in the right carotid artery, suggesting that elevated cIMT measurement angles may coincide with increased disease risk. This observation also points to a potential disparity in the impact of left versus right carotid thickening on PCAD. Collectively, these results advocate incorporating cIMT monitoring into primary prevention guidelines for cardiovascular diseases and highlight the potential of imaging-based biomarkers to identify high-risk individuals before the onset of clinical symptoms. Such an approach would offer a critical window for timely intervention to mitigate cardiovascular events in younger populations.

### 4.3. Unanswered questions and future research

There are several limitations to our study. Although multivariable Cox proportional hazards models adjusted for demographic characteristics, conventional cardiovascular risk factors, and socioeconomic indicators (detailed in Table 1), inherent selection bias in observational designs may compromise causal inference validity. Second, current international guidelines lack consensus on diagnostic thresholds for subclinical atherosclerosis, with the Mannheim criteria recommending <0.9mm*[42]* versus AHA consensus adopting <1.0mm*[16]*, our use of 1.0mm threshold based on majority cohort standards may introduce disease misclassification bias. Furthermore, the strict sex-specific age thresholds may have led to excessive case exclusion, with reduced subclinical atherosclerosis cases in the premature cohort potentially compromising statistical robustness. Finally, UK Biobank participants exhibited healthier baseline behaviors, limiting generalizability, although such selection bias typically preserves internal validity of exposure-disease associations.

## 5. Conclusion

Based on large-scale prospective cohort data from the UK Biobank, this study confirms a significant positive association between cIMT and the risk of both PCAD and LCAD. These findings highlight the potential of early intervention in subclinical atherosclerosis to reduce coronary artery disease risk. Moreover, even within conventional “safe thresholds”, cumulative cIMT exposure remains linked to a progressive risk gradient, underscoring the limitations of static threshold-based cardiovascular risk models. Future research should integrate dynamic cIMT trajectories and multi-omics data to develop predictive models and identify high-risk individuals for early-onset coronary artery disease, facilitating the discovery of mechanistically informed biomarkers for precision risk stratification.

## Data Availability

All data used in this study are publicly accessible from UK Biobank via their standard data access procedure at https://www.ukbiobank.acuk/. Code used in the current study is available from the authors upon request.

## ABBREVIATIONS ANDACRONYMS

PCAD: Premature coronary artery disease
LCAD: later-onset coronary artery disease
cIMT: carotid intima-media thickness
CTree: conditional inference tree
GBP: Great Britain Pound
SBP: Systolic blood pressures
DBP: Diastolic blood pressure
BMI: body mass index
SD: standard deviation
RCS: restricted cubic spline

## Acknowledgements

All authors have participated in the work and have reviewed and agree with the content of the article. None of the article contents are under consideration for publication in any other journal or have been published in any journal. No portion of the text has been copied from other material in the literature (unless in quotation marks, with citation). I am aware that it is the author’s responsibility to obtain permission for any figures or tables reproduced from any prior publications, and to cover fully any costs involved. Such permission must be obtained prior to final acceptance. We are indebted and thankful to all participants and staff from this prospective cohort study for their valuable contributions.

## Author contributions

J.L. and L.T. contributed to the conception or design of the work. M.G., M.Z., Y.Z., A.W. and J.L. contributed to the acquisition, analysis, or interpretation of data for the work. M.G. and J.L. drafted the manuscript. X.G. and L.T. critically revised the manuscript. The authors declare no competing interests. All authors gave final approval and agreed to be accountable for all aspects of work ensuring integrity and accuracy.

## Appendices

**Table 1.**
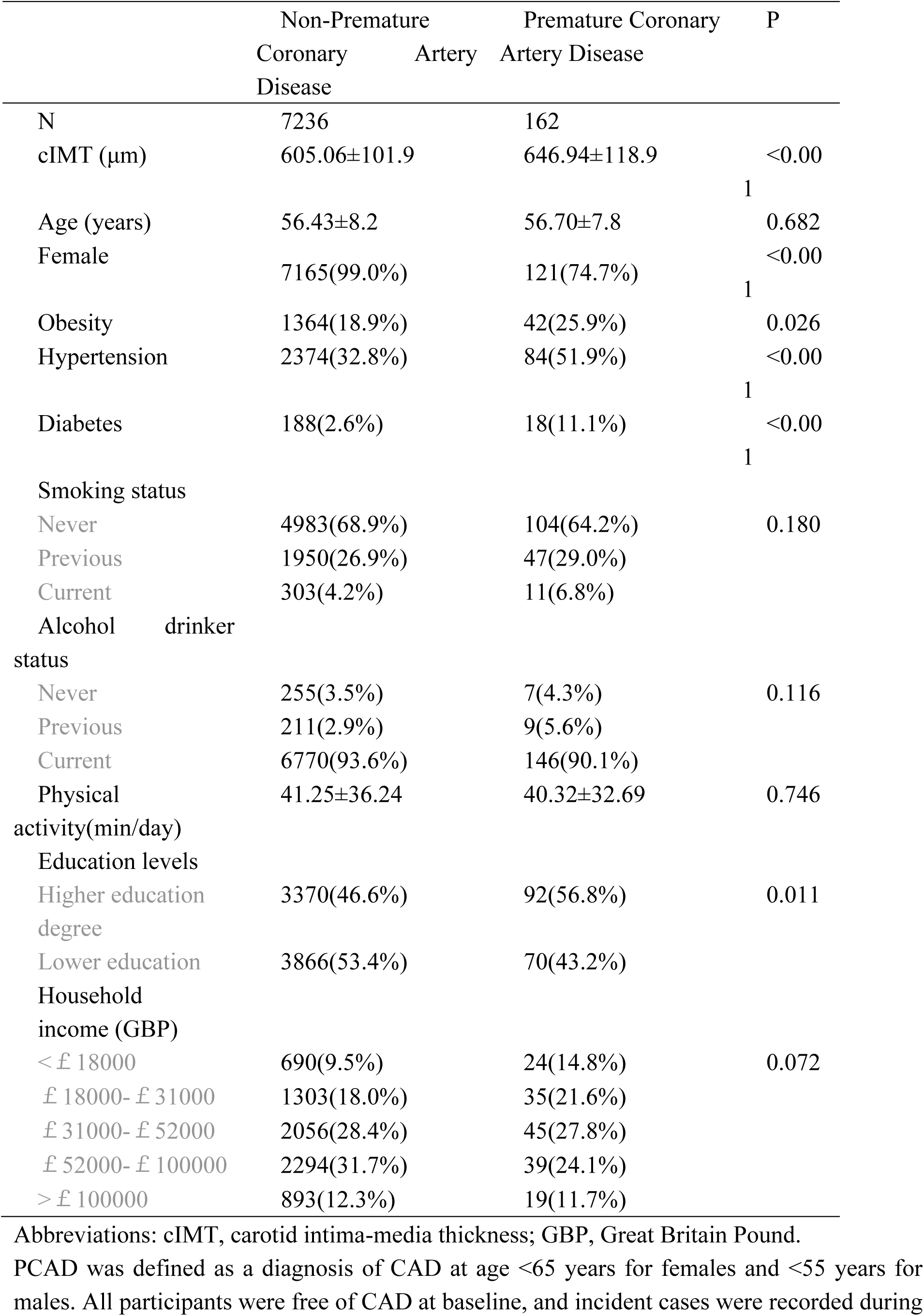
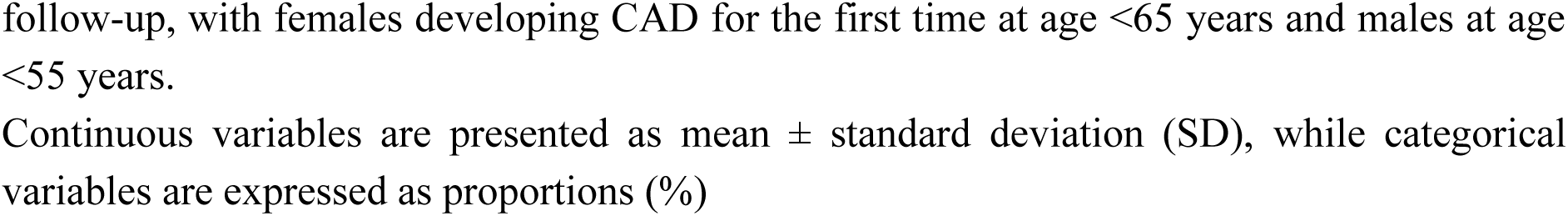
Baseline characteristics of PCAD cohort.

